# Evaluating biomedical feature fusion on machine learning’s predictability and interpretability of COVID-19 severity types

**DOI:** 10.1101/2024.04.04.24305295

**Authors:** Haleigh West-Page, Kevin McGoff, Harrison Latimer, Isaac Olufadewa, Shi Chen

## Abstract

**Background:** Accurately differentiating severe from non-severe COVID-19 clinical types is critical for the healthcare system to optimize workflow. Current techniques lack the ability to accurately predict COVID-19 patients’ clinical type, especially as SARS-CoV-2 continues to mutate. We explore predictability and interpretability of multiple state-of-the-art machine learning (ML) techniques trained and tested under different biomedical data types and COVID-19 variants.

**Methods:** Comprehensive patient-level data were collected from 362 patients (214 severe, 148 non-severe) with the original SARS-CoV-2 variant in 2020 and 1000 patients (500 severe, 500 non-severe) with the Omicron variant in 2022-2023. The data included 26 biochemical features from blood testing and 26 clinical features from patients’ clinical characteristics and medical history. Different ML techniques including penalized logistic regression (LR), random forest (RF), *k*-nearest neighbors (kNN), and support vector machines (SVM) were applied to build predictive models based on each data modality separately and together for each variant. Fifty randomized train-test-splits were conducted per scenario and performance results were recorded.

**Findings:** The fused (hybrid) characteristic modality yielded the highest mean area under the curve (AUC) achieving 0·915, while the biochemical modality alone and the clinical modality alone had AUCs of 0·862 and 0·818 respectively. All ML models performed similarly under different testing scenarios and were robust when cross-tested with original and Omicron variant patient data. Our models ranked elevated d-dimer (biochemical), elevated high sensitivity troponin I (biochemical), and age greater than 55 years (clinical) as the most predictive features of severe COVID-19.

**Interpretation:** ML is a powerful tool for predicting severe COVID-19 based on comprehensive individual patient-level data. Further, ML models trained on the biochemical and clinical modalities together witness enhanced predictive power. The improved performance of these ML models when trained and cross-tested with Omicron variant data supports the robustness of ML as a tool for clinical decision support.

**Funding:** U.S. Centers for Disease Control and Prevention (CDC)

**Research in Context:** *Evidence before this study:* We searched the PubMed database for publications investigating the use of machine learning (ML) in predicting severe COVID-19 types using patient-level data. We found studies published from the beginning of the COVID-19 pandemic in 2020 up to February 2023 using keywords such as “severe COVID-19”, “SARS-CoV-2”, “multimodal”, “machine learning”, “prediction”, and “data-driven.” The resulting studies were overall limited in scope, as they focused on single data modalities or uninterpretable models. Nearly all studies found only used patient data obtained from the outbreak of COVID-19 and lacked data from the later variants, such as Omicron. These limitations prevent identification of the data modalities and ML techniques most suitable for predicting severe types, as well as the generalizability of these models to multiple variants.

*Added value of this study:* We built end-to-end machine learning pipelines with a variety of ML techniques, data modalities (biochemical, clinical, and fusion), and SARS-CoV-2 variants (original and Omicron) to compare the predictive power of each model type. Our study shows these models have strong predictive power severe COVID-19 when trained on multiple modalities and robustness across different variants of the virus, with two models achieving an AUC > 0·90. We compared feature rankings of models trained with the different variants and found overall agreement that the following features are highly predictive of severe COVID-19: elevated coagulation markers (d-dimer), indicators for heart damage (hsCRP, hsTNI), and age >55 years.

*Implications of all the available evidence:* These findings result from a thorough analysis of the effect of data type, ML technique, and SARS-CoV-2 variant on the power to predict severe COVID-19. To our knowledge, no other work has provided analysis of the effect of these characteristics, particularly the SARS-CoV-2 variants, on the performance of ML models. This model yields a powerful framework for healthcare providers seeking clinical decision support tools for not only COVID-19, but many other viral respiratory illnesses. Our work demonstrates a need for further testing with larger datasets to confirm the benefits of biomedical feature fusion.

## Introduction

The COVID-19 pandemic caused by the SARS-CoV-2 virus has impacted healthcare systems everywhere. Since 2019, several major SARS-CoV-2 variants and sub-variants have manifested, with the Omicron variant being the most persistent since November 2021.^1^ A critical effect of the pandemic has been the sudden increased burden on healthcare facilities, mostly hospitals. The influx of severe COVID-19 patients overwhelms intensive care units which results in increased mortality,^2^ especially in regions with fewer health resources.^3,4^

In current clinical practice, patients with COVID-19 are typically classified as having severe illness by features such as shortness of breath, low oxygen saturation, and low PaO_2_/fraction of inspired oxygen. However, these few features cannot sufficiently distinguish between severe and non-severe types of patients with COVID-19, as some severe types may lack these or any symptoms upon admission.^5^ Without suitable medical intervention these severe types may progress quickly to a critical condition, resulting in a high risk of mortality.^6^ This uncertainty motivates a predictive method of patient types that is reliable and efficient, while also making use of alternative features. Early determination of patient types may enable healthcare professionals to improve their treatment plans and optimize facility resources.

In this study, we investigated the performance of various machine learning (ML) techniques for COVID-19 severity prediction, and then we evaluated feature modalities that provide the most accurate and reliable results. We trained ML models employing different techniques with patient-level biochemical and clinical feature modalities, both separately and together as a fusion modality. We applied logistic regression (LR), decision tree-based random forest (RF), *k*-nearest neighbors (kNN), and support vector machines (SVM), and evaluated their abilities to predict severe COVID-19. We developed these ML models from data collected from original and Omicron COVID-19 patients to investigate model robustness across different variants.

## Methods

### Data Collection

Our study uses two distinct datasets covering two time periods with distinct dominant viral variants. All patients were confirmed to be positive for COVID-19 by two independent quantitative reverse transcriptase-polymerase chain reaction (qRT-PCR) tests before inclusion in this study. The first set includes 362 patients infected with the original SARS-CoV-2 variant upon admission to Wuhan Union Hospital in China from January to March 2020. This dataset was previously described and analyzed by Chen et al.^5^ and serves as a comparative baseline in this study. Among these 362 patients, 148 were in severe condition according to guidelines established by the National Health Commission of China and the American Thoracic Society,^7,8^ while the remaining 214 were designated non-severe types. Severe types were categorized by meeting at least one of the following criteria: (i) respiratory rate >30 breaths per minute; (ii) oxygen saturation <93% at rest; or (iii) PaO/fraction of inspired oxygen <300 mm Hg (40 kPa). As this dataset contains data from patients infected by the original SARS-CoV-2 variant, this set is referred to as “original” in the rest of this study. The second dataset consists of 1000 patients admitted to Wuhan Union Hospital in China from December 2022 to January 2023, during which time patients were diagnosed with the SARS-CoV-2 Omicron variant. Using the same guidelines outlined earlier, 500 of these patients were severe COVID-19 type, while the other 500 were non-severe type.

All patients were comprehensively evaluated before being admitted to the hospitals. Their fully de-identified, anonymous biomedical data were extracted from the electronic health record system. All participants were informed about the study, agreed to participate, and signed written consent. An institutional review board (IRB) application was submitted and approved by the Wuhan Union Hospital, Tongji College of Medicine, Huazhong University of Science and Technology (IRB approval #IEC-J-345), where the data were collected.

The de-identified patient information comprised two main modalities of biomedical features. The first feature modality had 26 distinct laboratory testing features from blood tests, most of which were continuous values of the readings. The specifics of these tests are reported in detail in our prior study.^5^ We refer to this feature modality as “biochemical” hereinafter. The second is a total of 26 features of one-hot encoded binary values indicating the presence of pre-existing conditions, comorbidities, symptoms, and other key risk factors such as demographic information. This modality was referred to as “clinical” features. A complete description of the features across these two modalities is present in the supplementary materials of our prior study.^5^ Together, features from both modalities were appended into a single corpus of de-identified patient data with 52 multimodal features. This was referred to as the “fusion” set, as it fused across continuous, real-valued biochemical feature modality and binary clinical feature modality. We note that the specific features of respiratory rate, oxygen saturation, and fraction of inspired oxygen were excluded from our predictive feature list as they were the original clinical standard to determine COVID-19 severity.

### ML Pipeline Development, Validation and Interpretation

We developed, evaluated, and compared the performance of several state-of-the-art ML classification techniques, including RF, kNN, and SVM. To acknowledge LR’s popularity in the field,^9–15^ we included it as a benchmark method. All ML techniques were implemented as supervised binary classification problems. Utilizing Python 3·10 and a variety of ML-related libraries from Sci-Kit Learn,^16^ we developed an end-to-end ML framework for each of the four classifiers to predict COVID-19 severity types from patients’ clinical, biochemical, and fused feature modalities. A full list of packages is included in the supplementary materials.

The non-severe and severe COVID-19 types were labeled as 0 and 1, respectively. Each classifier was constructed to predict the outcome (0 or 1) based on the input features provided. For a given dataset, the corpus was randomly partitioned into training and hold-out testing sets by an 80% to 20% split, respectively. The biochemistry feature data were then preprocessed by a standard scaler prior to training to ensure consistency across different features. During the training step, a grid search method of hyperparameter tuning was utilized to maximize the ML model’s performance. The resulting optimal hyperparameters for each classifier were detailed in the supplementary materials. Upon completion, the models with trained hyperparameters were then applied to the hold-out data for testing. This process was repeated 50 times to generate different random training-testing splits, which avoids overfitting^17^ and establishes an average performance.

ML classifiers’ performances were evaluated by the receiver operating characteristic (ROC) curve and computing the area under the curve (AUC) of ROC. In this study, AUC was selected as the main performance metric (as opposed to F-measure or accuracy) because AUC has been proven to be more reliable than the other metrics.^18^ For both the original and Omicron variant data sets, we evaluated ML classifier performance of training and testing on each modality separately and fused.

Each data set (binary clinical feature modality alone, continuous biochemical feature modality alone, and fused modality that incorporates both feature modalities) underwent the pipeline defined earlier. To evaluate the robustness of the developed ML classifiers, we swapped and cross tested with testing data from the other variant. For example, models trained on the original SARS-CoV-2 variant data were also tested with Omicron variant data, and *vice versa*: this process was mirrored for classifiers trained on the Omicron variant data. For cross testing, the testing data was standardized according to the scaling scheme from the classifier’s training data. During cross testing, the entire corpus was used as a hold-out testing set, since the classifiers were only trained with one variant and never trained with the other variant’s data. The cross-set testing was evaluated for its performance exactly as the same-set testing.

One of the advantages of certain ML classification techniques is their interpretability in addition to performance. Of the classifiers that were developed in this study, we gathered insights from LR and RF classifiers. The resulting feature coefficient vector obtained from training LR indicated what the classifier “learned” from the data. This appears as the largest absolute coefficient corresponding to the most influencing feature in predicting the severe COVID-19 type.^19^ Pertaining to RF, feature importance was quantified and ranked by the feature’s mean decrease in Gini impurity,^20^ which is commonly used in feature selection tasks.^21^ During RF’s training, these feature importances were computed using Sci-Kit Learn’s feature_importances_ package.^20^ By averaging the feature rankings (i.e., importance in predicting severe COVID-19 type) over 50 runs, we compared feature importance identified by LR versus RF, as well as different feature importances between the original and Omicron variant data. These comprehensive investigations enable us to validate findings from the ML classifiers by cross-checking results with other studies performing traditional statistical studies aimed at identifying predictive features of the severe COVID-19 type.

**Figure 1:**
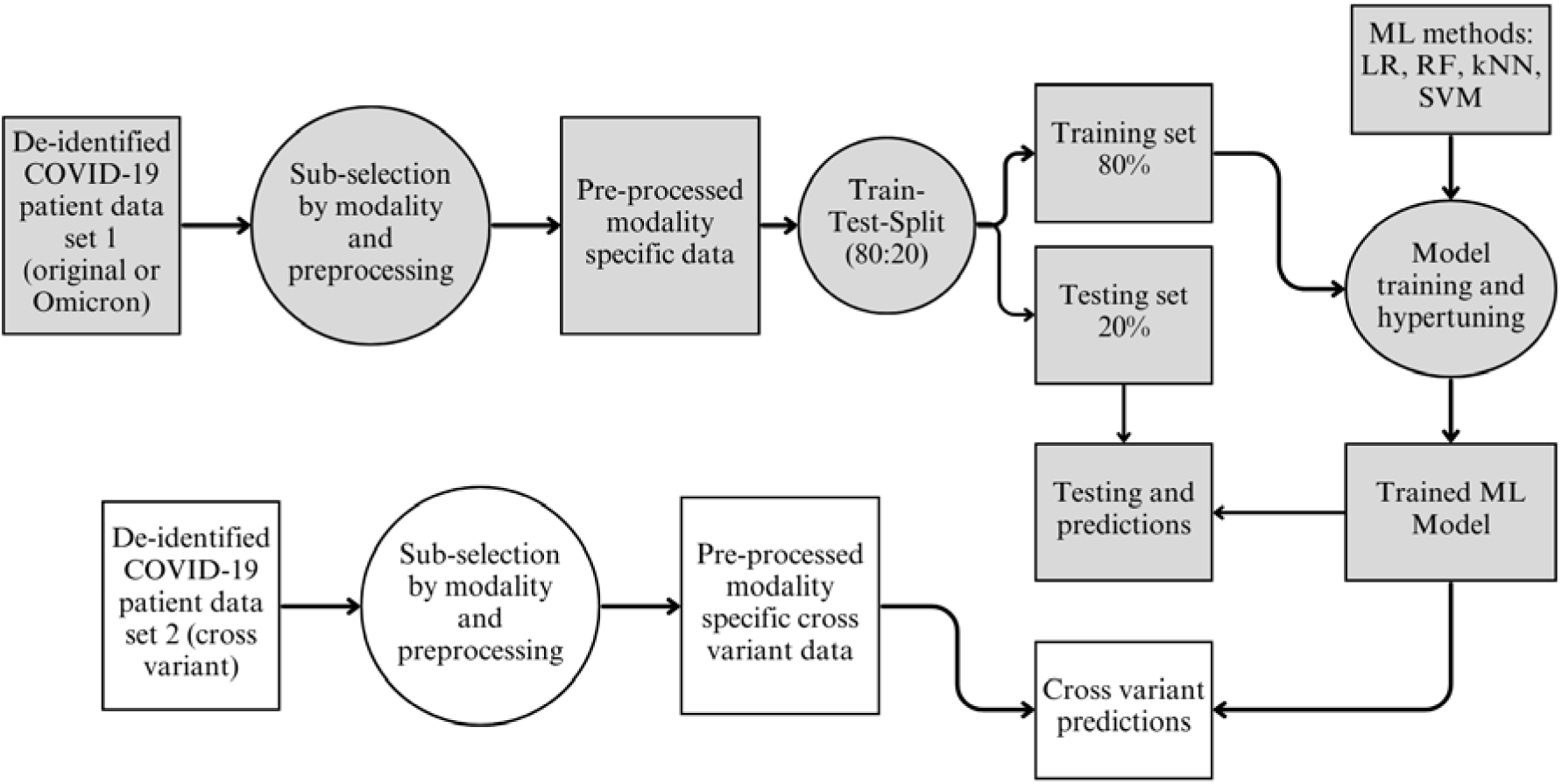
Machine learning model pipeline design. De-identified patient biomedical data were collected from Wuhan Union Hospital from January to March 2020 (original COVID-19 patients n=362) and December 2022 to January 2023 (Omicron variant patients n=1000). The data were grouped by their feature modality: biochemical, clinical or fusion. An 80:20 split was performed to generate training and testing sets, respectively. Four classifiers were developed and evaluated for their performance: LR=logistic regression, RF=random forest, kNN=k-nearest neighbors, and SVM=support vector machine. Upon hyperparameter tuning, each classifier was validated with the same-variant hold-out set, as well as the preprocessed cross-variant set.

## Results

### ML Classifiers Performance

Upon running each ML classifier pipeline for 50 independent repetitions, the average AUC value was calculated (Table 1). We validated that the computed average AUC was equal to the AUC of the composite ROC curve, hence there is no need for their distinction. These values were tabulated according to which SARS-CoV-2 variant dataset and modality were used for training each ML classifier. The standard deviations (SD) of the AUC did not exceed 0·06 across all testing scenarios. A summary of the standard deviations is found in the supplementary materials. Our tuned ML classifiers demonstrated overall high accuracies in predicting severe COVID-19. In general, ML models trained from Omicron variant data performed the best across all scenarios. The highest AUC among classifiers trained on Omicron data was 0·915, compared to the original’s highest at 0·827 (Table 1). Performance of different ML classifiers trained on the same dataset showed minor differences. All ML classifiers developed from either original or Omicron data were almost indistinguishably accurate when tested on the Omicron data. For each ML classifier, ROC plots were generated for the same-variant and cross-variant testings, yielding a total of 4 training-testing combinations (original-original, Omicron-Omicron, original-Omicron, and Omicron-original, where the latter two were cross-variant testings). Each ROC plot visualizes comparisons between the three feature modalities (clinical alone, biochemical alone, and fusion). All 16 plots are presented in the supplemental materials. For brevity, we discuss four graphs from RF in Figures 2(a)-(d). Each graph shows the composite ROC plot over 50 independent repetitions for each modality. The shaded regions denote one standard deviation from the mean of the true positive rate (TPR). The red, green, and blue lines represent the performance of models with biochemical, clinical, and fusion feature modalities, respectively.

**Figure 2:**
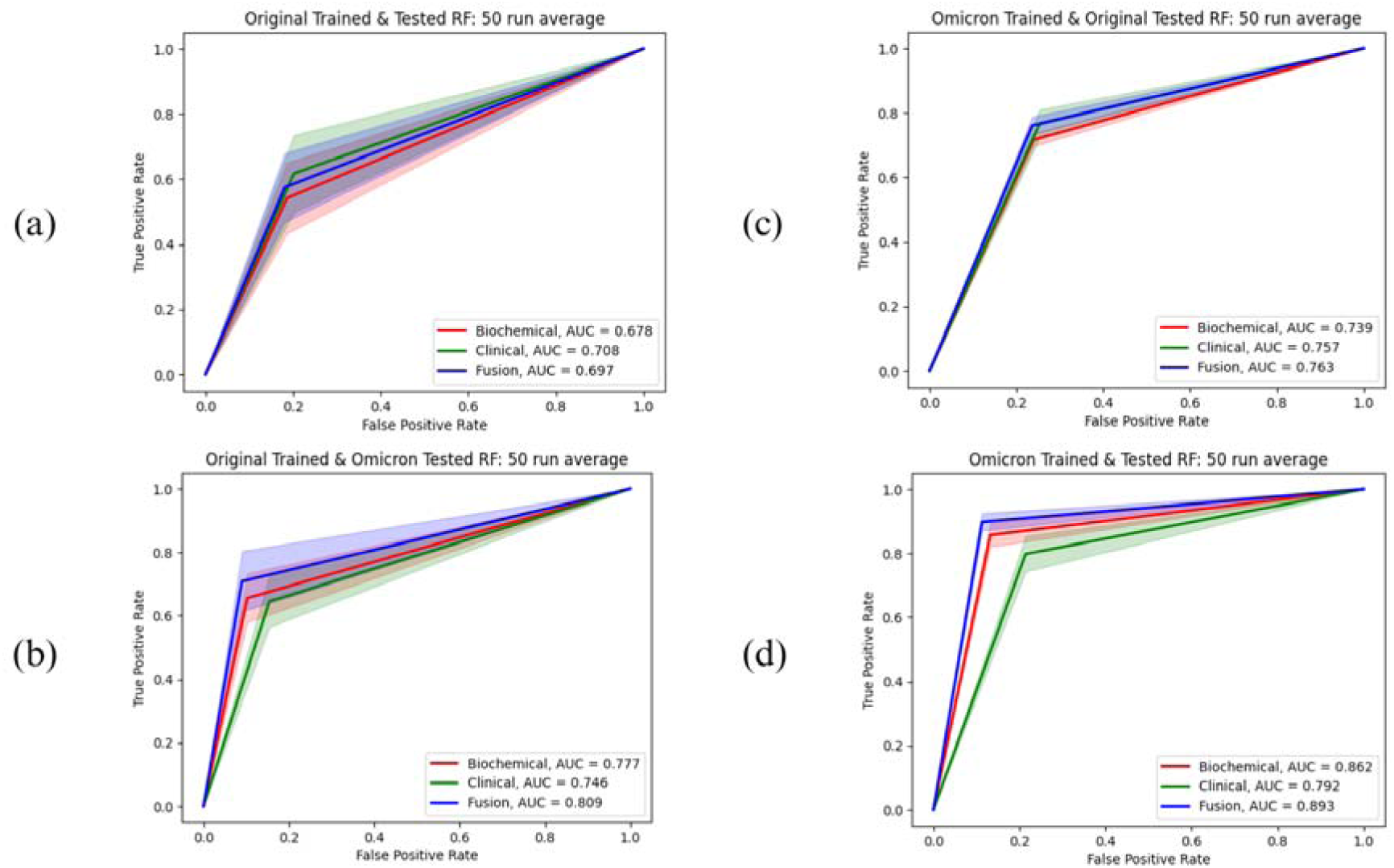
ROC curve plots for random forest (RF) This figure shows the mean ROC curves plotted for each of the four training-testing combinations. Each figure contains the ROC curve of each data modality (biochemical-red, clinical-green, and fusion-blue.) Panel (a) plots the curve of classifiers trained and tested on original data only (i.e., original-original combination). Panel (b) plots the curve of classifiers of the original-Omicron combination. Panel (c) plots the curve of classifiers of the Omicron-original combination. Panel (d) plots the curve of classifiers of the Omicron-Omicron combination.

**Table 1:**
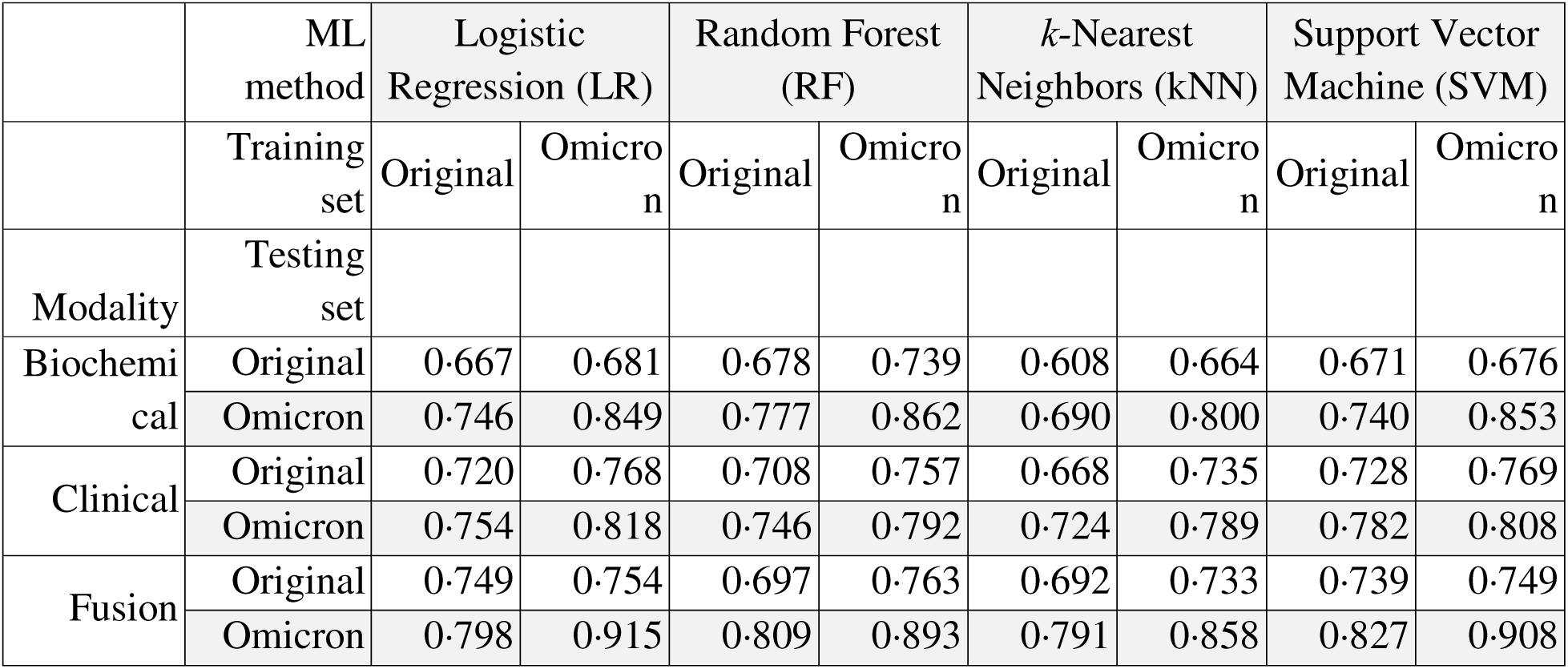
Area under the ROC curve (AUC) values. This table displays the mean AUC values of each machine learning classification technique when applied to various training and testing combinations over 50 random independent splits. For each classifier, a model was developed, trained, and tuned with either original or Omicron COVID-19 patient data. The developed classifier was tested on hold-out data from both the same variant and the cross-variant data. Values are separated by training-testing combinations, as well as feature modality for training.

Classifiers trained with the fusion feature modality generally demonstrated higher predictive power than either biochemical or clinical feature modality alone. The clinical feature modality performed slightly worse than the fusion modality, while biochemical feature modality alone had the least predictive power. Regardless of ML classification technique, models trained and tested with original variant data experienced the largest variation in their performance. We note that this combination also had the lowest performance of the four training-testing combinations. Classifiers trained with original data had improved performance when cross-testing with Omicron data.

### Feature Importance Ranking

During each replication of the ML classifier, the feature coefficient vectors from the tuned LR classifier and the mean decrease of Gini impurity score vectors from the RF classifier were recorded and averaged after 50 replications. The averaged coefficients were then used to identify important features that differentiate severe from non-severe COVID-19 and to evaluate how different training data (original or Omicron variant) and feature modalities influence the feature rankings. Note that LR’s associated coefficient vector has real values, while RF’s reduction of Gini importance is interpreted as probabilities in [0, 1].

In this study, feature importances were determined by the fusion modality, as feature fusion demonstrated the highest predictive power for severe COVID-19. Regardless of ML classifiers (LR and RF) or SARS-CoV-2 variant, features such as d-dimer (DD; biochemical modality), high sensitivity Troponin I (hsTNI; biochemical), and age > 55 years (OLD; clinical modality) were consistently ranked in the top five most predictive features for COVID-19 severity. Features that often appeared among the top ten also include high sensitivity C-reactive protein (hsCRP; biochemical) and hypertension (HYP; clinical).

There were also some disagreements in feature rankings between the two techniques. For instance, LR suggested a history of chronic obstructive pulmonary disease (CPD; clinical) as an important predictive feature of the severe COVID-19 when trained on both the original and Omicron variant data, but CPD was not identified as a top feature in RF. Conversely, only RF identified elevated lymphocytes (LY; biochemical), ferritin (FERR; biochemical), and interleukin-6 (IL-6; biochemical) as important features regardless of SARS-CoV-2 variants.

By comparing the feature rankings across variants, LR trained on the original variant data identified low-, mid- and high-grade fever (LOD, MDF, HIF; clinical) all among the top 10 most predictive features, while its counterpart trained on Omicron variant data identified elevated procalcitonin (PCT; biochemical), percent of neutrophil (NE.1; biochemical) and white blood cell (WBC; biochemical) as the most predictive features. Such discrepancies in feature rankings were not observed in results from RF classifiers trained on different variants’ datasets.

Lastly, there were some differences in the range of feature importance (quantified by coefficients in LR and Gini impurity scores in RF) across the two variants. LR’s feature coefficients on average fell in the range [-0·95, 2·30] for the original variant, whereas the range was [-0·70, 2·85] for the Omicron variant. Mean decreases of Gini impurity were in the range of [0, 0·10] and [0, 0·12] for the original and Omicron variants, respectively.

**Figure 3:**
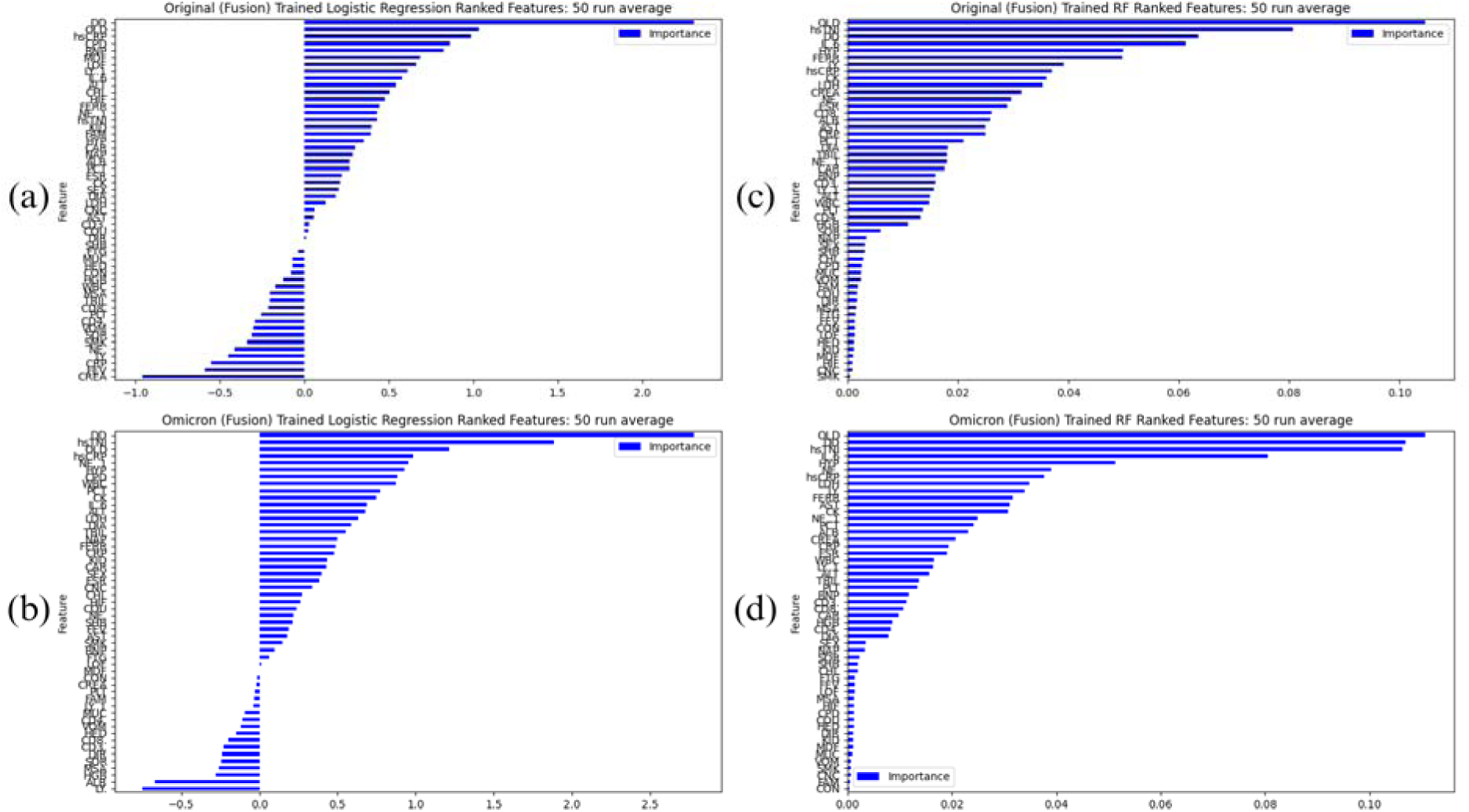
Feature rankings. This figure displays the mean feature rankings from LR classifiers trained on the original variant dataset in panel (a), LR on Omicron variant in panel (b), RF on original variant in panel (c), and RF on Omicron variant in panel (d). Feature rankings are determined by the coefficients of the feature weight vector in LR, and Gini impurity scores in RF.

## Discussion

In this study, we evaluated the predictive power of multiple ML techniques when utilizing different feature modalities. We also found differences in model performance and interpretations across different SARS-CoV-2 variants. Overall, we found ML to be a powerful tool for predicting COVID-19 severity based on comprehensive individual patient-level data. More importantly, we discovered that fusion of the biochemical and clinical modalities enhanced the predictive power of all types of ML models evaluated in this study. Models trained on multiple feature modalities have yielded the best performance in nearly every performance metric across all testing sets. These multimodal features are accessible by healthcare systems, especially with wide adoption of electronic health record systems. Results can be obtained efficiently from these systems, allowing the predictive ML model to be a fast and reliable clinical decision support tool to identify patients with high risk of severe COVID-19.^22^

The similarity of performance between the four ML techniques evaluated in this study suggests that the specific choice of modeling technique is not important for the task of predicting severe COVID-19 types. In general, LR, RF, and SVMs all showed relatively strong performance with their highest AUC scores being 0·915, 0·893 and 0·908, respectively. The kNN model is the relative lowest, with its highest AUC being 0·858. If model interpretability is important to the clinical decision support of these ML models, then LR and RF should be considered. The LR model offers the analyst information on which features are positively and negatively associated with the risk of severe COVID-19. However, LR is susceptible to multicollinearity between different features.^23^ The RF model, on the other hand, is more resilient to the multicollinearity issue in the input data.^24^ The RF model’s reliability and robustness are shown in the findings of this study, and further supported by other similar studies.^5,12^

Upon validation, this study was among the first to use ML to identify critical biomedical features with the most predictive power of differentiating severe COVID-19 patients across different dominant variant phases. The feature rankings provided by LR and RF are important for clinical decision making and provide clues to COVID-19 pathology. Our study indicates that elevated biomarkers such as D-dimer for coagulation, as well as hsTNI and hsCRP as indicators for heart damage, are strongly associated with severe COVID-19. Other works have also shown that cardiovascular injury due to COVID-19 is highly associated with severe disease and adverse patient outcomes.^25^ Higher D-dimer is associated with higher risk of progressing to severe stage.^26^ Our findings also suggest that patients’ clinical information such as being 55 years or older, or having pre-existing conditions such as hypertension and COPD, could significantly increase the risk of progressing to severe COVID-19. Other studies also confirm age and hypertension as major risk factors for severe COVID-19.^27–29^

When comparing important features between patients infected by original and Omicron variants, we have identified an increase in the feature weight vector and Gini impurity values, which have not been reported before. This finding suggests that COVID-19 severity became more predictable in more recent variants. We speculate that patient-level data may have higher quality in the Omicron wave than the original variant. This might also explain the higher variability and lower performance of models trained and tested on the original SARS-CoV-2 patient data.

This study contains a variety of limitations. One hindrance to the generalizability of this framework is the lack of variation in data samples. All patient data were taken at the individual’s time of admission to one healthcare facility, the Wuhan Union Hospital, and most patients were of the Han Chinese ethnicity group. This could result in sampling bias and the results should be further validated with larger scale multi-center studies.^30^ For this framework to be more robust, incorporation of larger datasets across more demographic groups would be necessary.

Another limitation is that we were not able to evaluate ML model predictability for other major SARS-CoV-2 variants, such as Alpha and Delta. Retrospective studies are needed to comprehensively evaluate the robustness of the developed ML models across different phases of COVID-19 with different dominant variants and subvariants.

There is a variety of future research directions suggested by this study. Notwithstanding improvements made on the limitations previously discussed, we acknowledge the existence of other emerging ML techniques worthy of investigation. Additionally, given the tentative promise of LR as a predictive tool for COVID-19 clinical decision support, other regression techniques such as the Lasso or Ridge regressions may be useful.

Further explorations shed light on the predictive power of ML techniques from individual-level data collected from patients with other respiratory illnesses. This is especially useful to healthcare systems inundated with patients infected with influenza or respiratory syncytial virus (RSV), to name a few. Using similar ML techniques and leveraging the power of transfer learning, our developed ML pipeline can be further applied to other diseases with similar underlying datasets (e.g., clinical and biochemical). Connecting back with the goal of aiding healthcare resource optimization, a potential application of this work is to simulate burdens on the health system of an unexpected inflow of patients, some of whom are severe patients and therefore need intensive care.

Another potential direction is the incorporation of more data modalities, such as patient-level medical imaging (including X Ray and CT scans) and multi-omics data. Due to the higher dimensionality of imaging modality in relation to the biochemical and clinical modalities, more advanced ML techniques such as a deep convolutional neural network (CNN) would need to be applied to handle such modalities.

## Contributors

SC conceptualized this study and the data. HW and SC designed the experiments and model structure. HW, SC, KM, HL and IO analyzed, interpreted and validated the results. HW drafted the manuscript. HW, SC, KM, HL and IO edited and revised the manuscript. HW, SC, KM, HL and IO had full access to the data used in this study and accept responsibility to submit for publication.

## Supporting information

Supplemental materials including packaged used, tuned hyperparameters, and complete list of ROC curves, feature importances

## Data Availability

All data produced are available online at

https://github.com/hnwestpage/Fusion-ML-COVID-19

## Data sharing

An institutional review board (IRB) application was submitted and approved by the Wuhan Union Hospital, Tongji College of Medicine, Huazhong University of Science and Technology (IRB approval #IEC-J-345), where the data were collected. This data and the code created for this study are available by visiting the GitHub repository here: https://github.com/hnwestpage/Fusion-ML-COVID-19

## Acknowledgements

The project described was supported by cooperative agreement (U01CK000677) from CDC. Its contents are solely the responsibility of the authors and does not necessarily represent the official views of CDC. KM acknowledges the support of NSF grants DMS-1847144 and DMS-2113676.

