## Supplemental materials including packaged used, tuned hyperparameters, and complete list of ROC curves, feature importances for "Evaluating biomedical feature fusion on machine learning’s predictability and interpretability of COVID-19 severity types"

| **Table 1: Sci-Kit Learn Packages** | |
| --- | --- |
| **Package Name** | **Purpose in code** |
| csv | Allows data retrieval from .csv files |
| pandas | Transform data into Pandas dataframes |
| numpy | Mathematical computations |
| matplotlib | Generating graphical displays |
| sklearn.preprocessing | Utilization of data scalars |
| sklearn.model_selection | Utilization of train_test_split algorithm and GridSearchCV |
| sklearn.linear_model | Initializes Logistic regression |
| sklearn.ensemble | Initializes RandomForestClassifier |
| sklearn.neighbors | Initializes KNeighborsClassifier |
| sklearn.svm | Initializes SVM |

| **Table 2: Model Hyperparameters** | | | |
| --- | --- | --- | --- |
| **Logistic Regression trained on original variant** | | | |
| Hyperparameters | Biochemical modality | Clinical Modality | Fused Modality |
| penalty | None | l1/l2 | l1 |
| **Logistic Regression trained on Omicron variant** | | | |
| Hyperparameters | Biochemical modality | Clinical Modality | Fused Modality |
| penalty | None | None/l1 | None/l1 |
| **Random Forest trained on original variant** | | | |
| Hyperparameters | Biochemical modality | Clinical Modality | Fused Modality |
| N_estimators (mean) | 8 | 7 | 7 |
| Max_depth (mean) | 5 | 5 | 5 |
| criterion | log_loss | entropy/log_loss | gini/log_loss |
| **Random Forest trained on Omicron variant** | | | |
| Hyperparameters | Biochemical modality | Clinical Modality | Fused Modality |
| n_estimators (mean) | 9 | 8 | 9 |
| max_depth (mean) | 8 | 6 | 6 |
| criterion | log_loss | gini | log_loss/gini |
| ***k*-Nearest Neighbors trained on original variant** | | | |
| Hyperparameters | Biochemical modality | Clinical Modality | Fused Modality |
| n_neighbors | 8* | 9 | 10 |
| weights | No preference | No preference | Uniform |
| metric | cosine | cosine | cosine |
| ***k*-Nearest Neighbors trained on Omicron variant** | | | |
| Hyperparameters | Biochemical modality | Clinical Modality | Fused Modality |
| n_neighbors | 8 | 9 | 10 |
| weights | distance | distance | distance |
| metric | cosine | cosine | cosine |
| **Support Vector Machine trained on original variant** | | | |
| Hyperparameters | Biochemical modality | Clinical Modality | Fused Modality |
| kernel | linear | sigmoid | sigmoid |
| **Support Vector Machine trained on Omicron variant** | | | |
| Hyperparameters | Biochemical modality | Clinical Modality | Fused Modality |
| kernel | linear | sigmoid | linear/rbf |

*Average was 8 neighbors, but median and mode were 9

| **Table 3: Results with standard deviations** | | | | | | | | | |
| --- | --- | --- | --- | --- | --- | --- | --- | --- | --- |
|  | ML method | Logistic Regression (LR) | | Random Forest (RF) | | *k*-Nearest Neighbors (kNN) | | Support Vector Machine (SVM) | |
|  | Training set | Original | Omicron | Original | Omicron | Original | Omicron | Original | Omicron |
| Modality | Testing set |  |  |  |  |  |  |  |  |
| Biochemical | Original | 0.667 (0.044) | 0.681 (0.007) | 0.678 (0.057) | 0.739 (0.012) | 0.608 (0.057) | 0.664 (0.010) | 0.671 (0.045) | 0.676 (0.007) |
|  | Omicron | 0.746 (0.026) | 0.849 (0.022) | 0.777 (0.029) | 0.862 (0.027) | 0.69 (0.034) | 0.800 (0.023) | 0.740 (0.033) | 0.853 (0.022) |
| Clinical | Original | 0.720 (0.061) | 0.768 (0.006) | 0.708 (0.054) | 0.757 (0.012) | 0.668 (0.060) | 0.735 (0.011) | 0.728 (0.043) | 0.769 (0.007) |
|  | Omicron | 0.754 (0.057) | 0.818 (0.023) | 0.746 (0.024) | 0.792 (0.030) | 0.724 (0.015) | 0.789 (0.024) | 0.782 (0.031) | 0.808 (0.024) |
| Fusion | Original | 0.749 (0.051) | 0.754 (0.007) | 0.697 (0.055) | 0.763 (0.014) | 0.692 (0.056) | 0.733 (0.008) | 0.739 (0.044) | 0.749 (0.009) |
|  | Omicron | 0.798 (0.058) | 0.915 (0.021) | 0.809 (0.043) | 0.893 (0.019) | 0.791 (0.027) | 0.858 (0.023) | 0.827 (0.044) | 0.908 (0.020) |

**Logistic Regression ROC Plots and Feature Rankings**

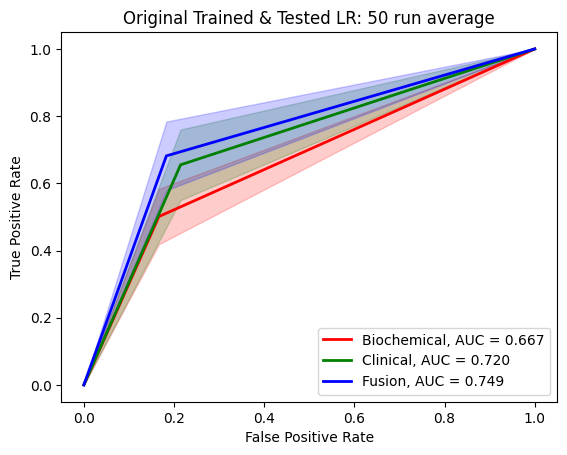

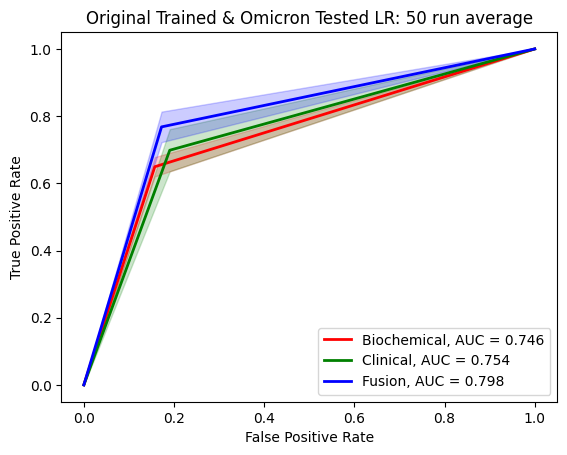

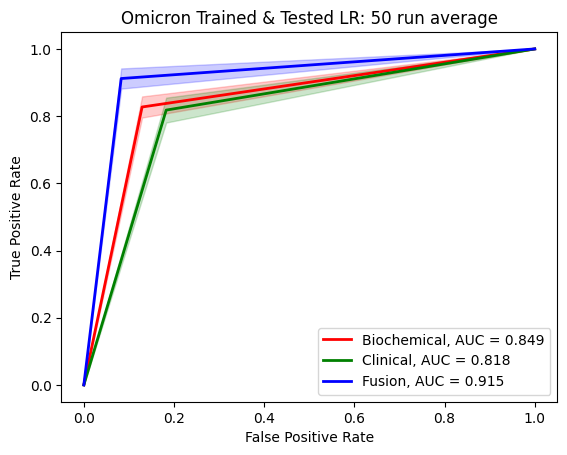

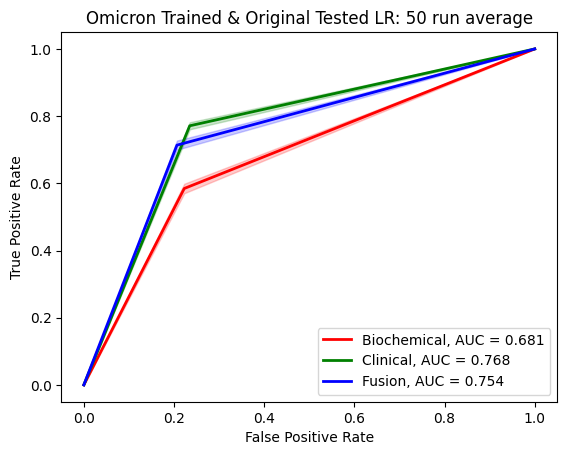

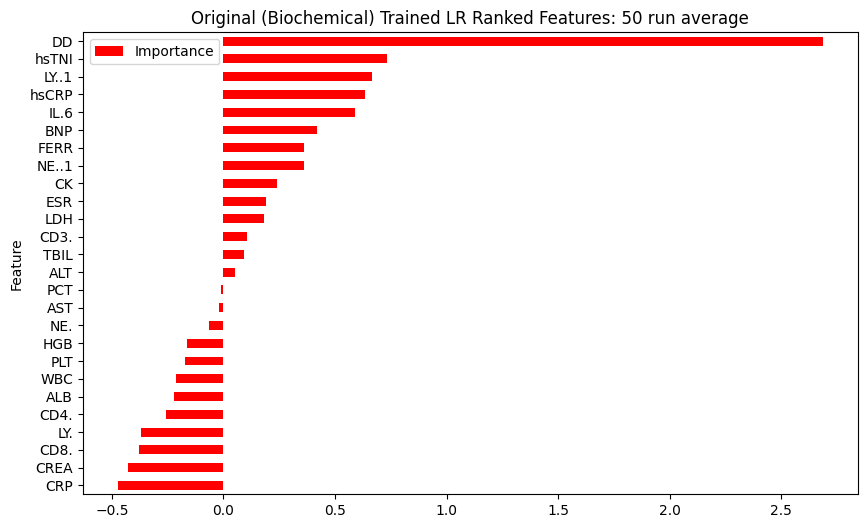

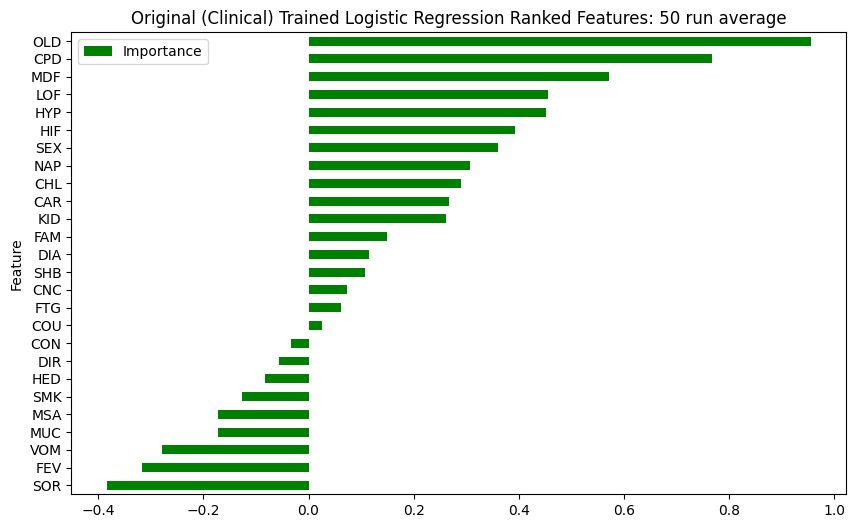

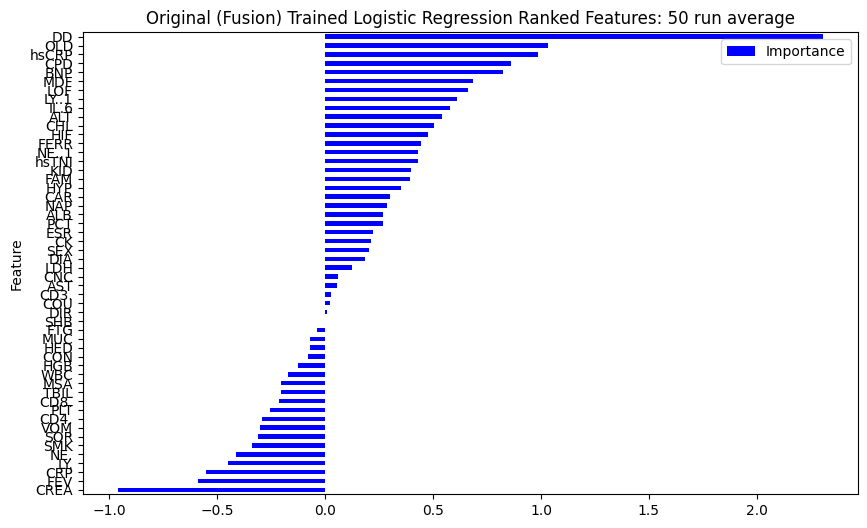

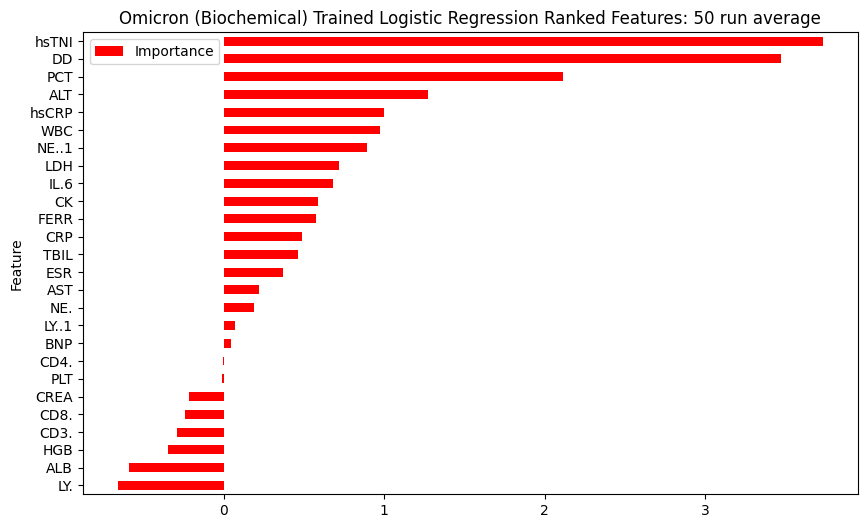

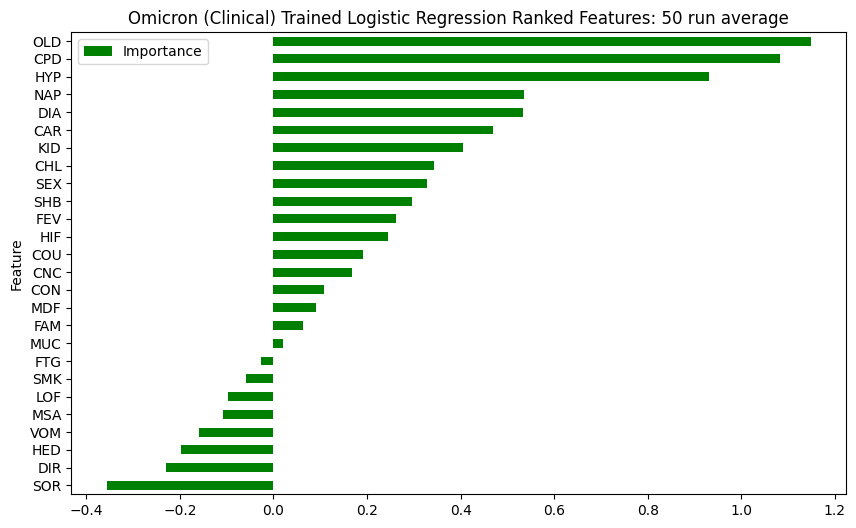

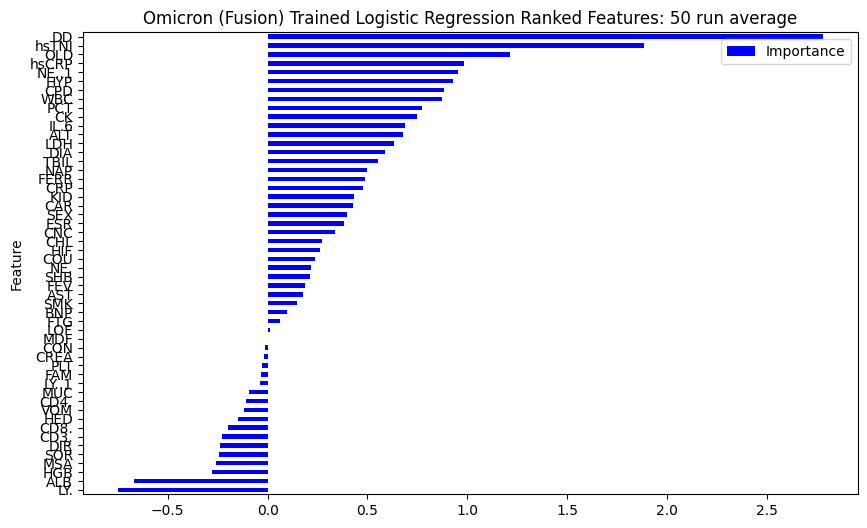

**Random Forest ROC Plots and Feature Ranking**

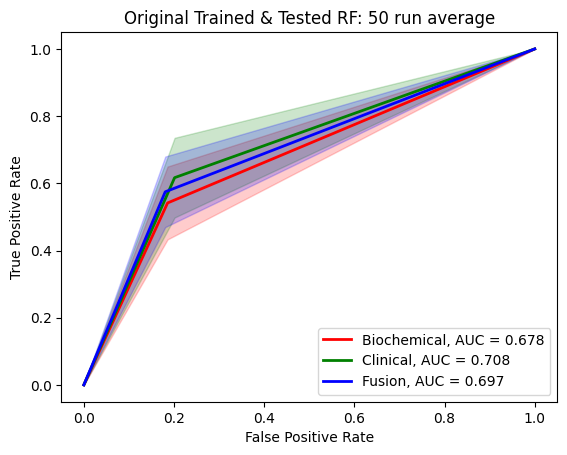

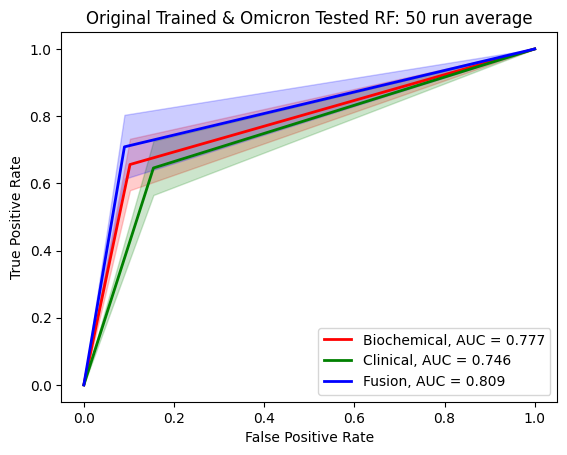

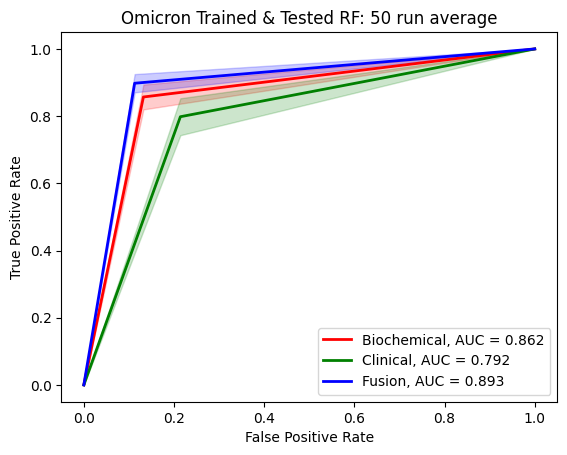

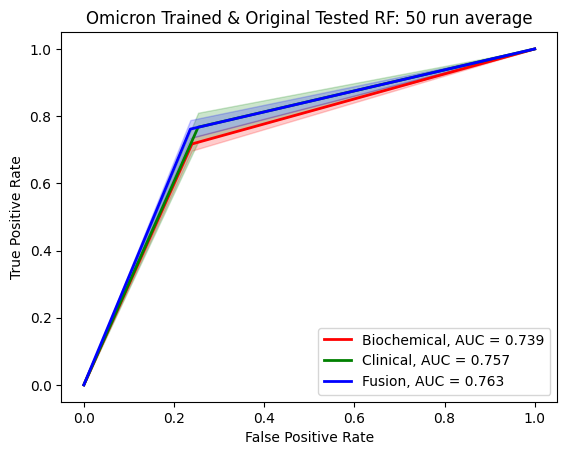

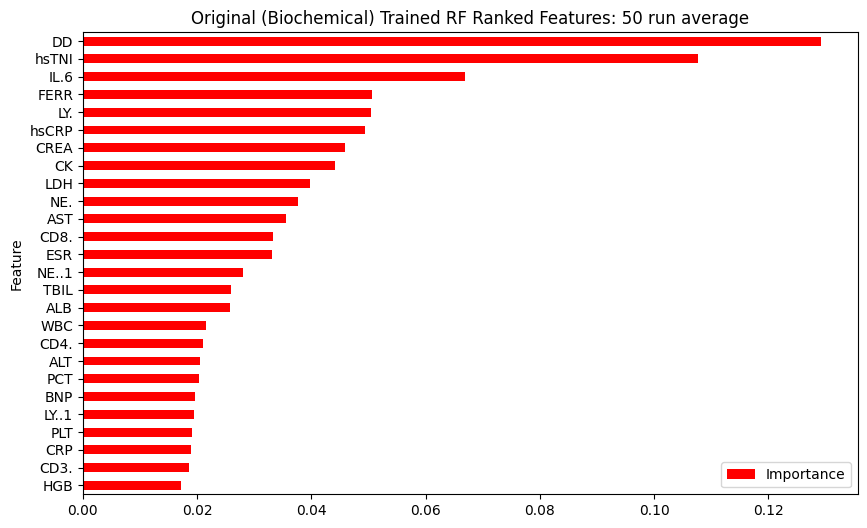

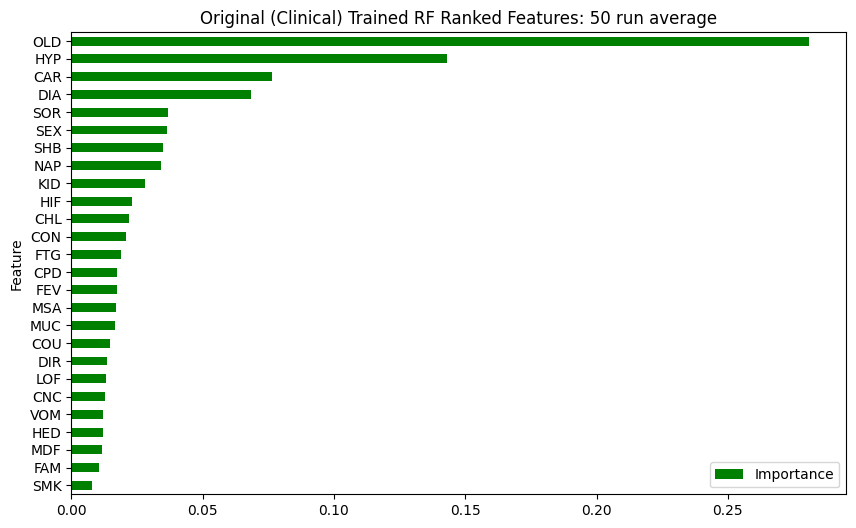

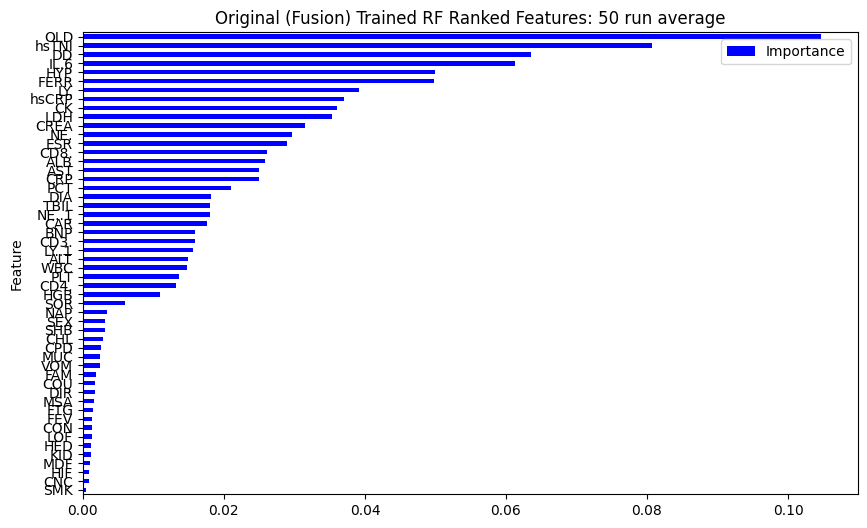

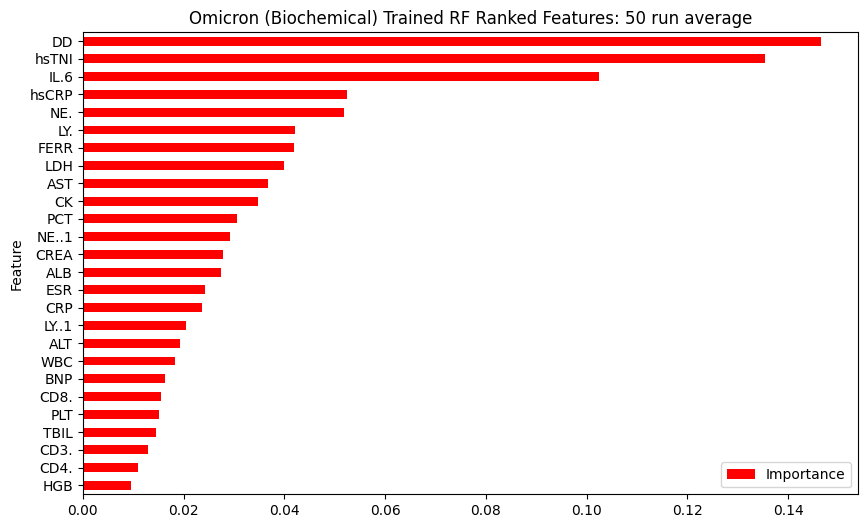

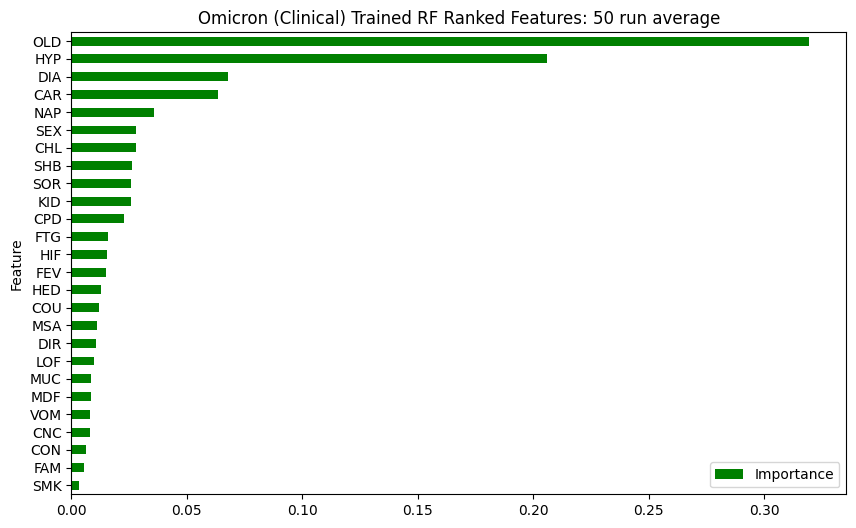

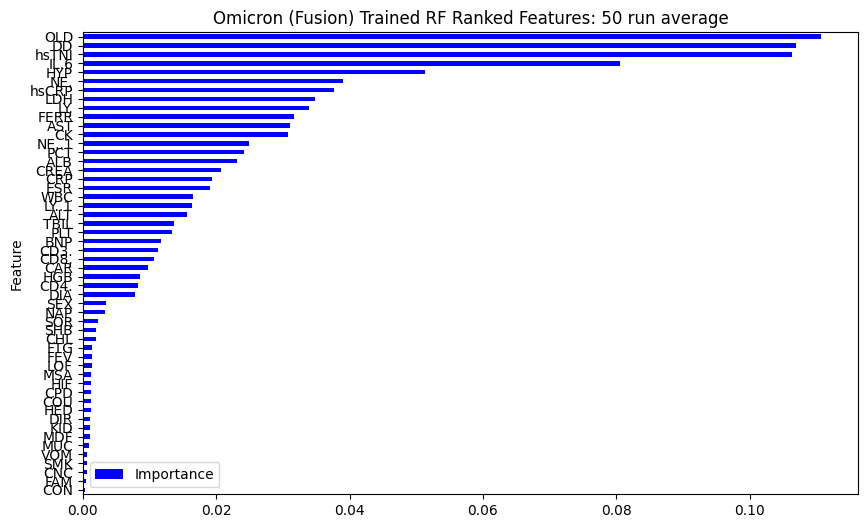

**K Nearest Neighbors ROC Plots**

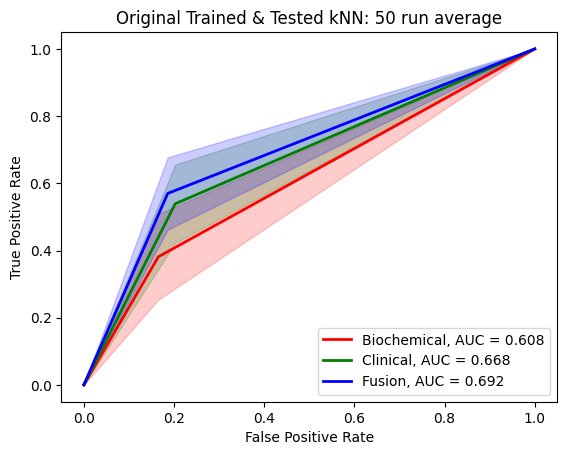

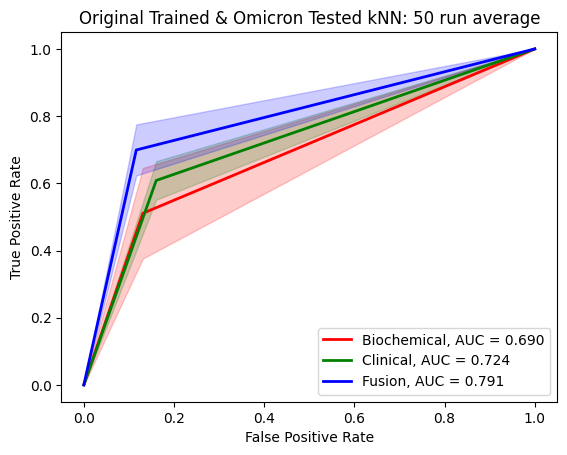

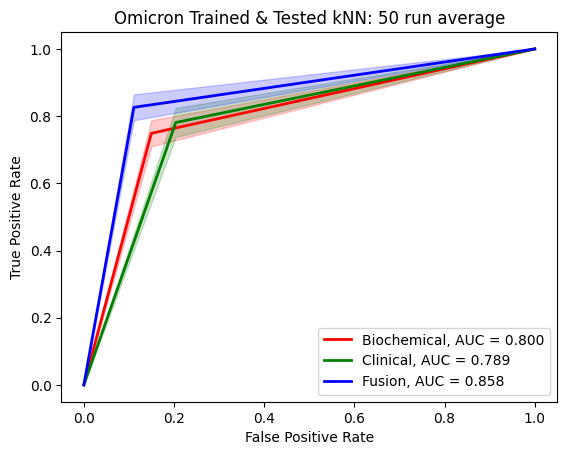

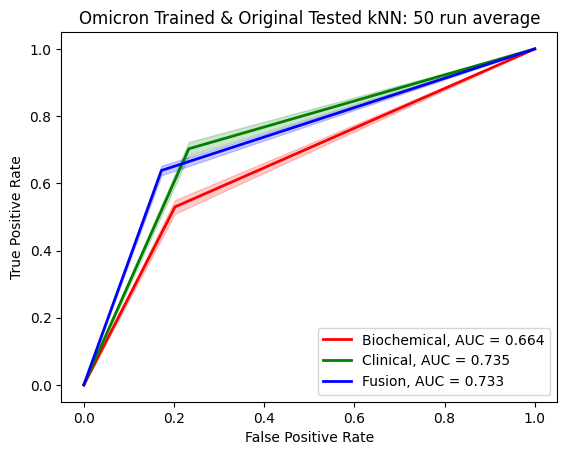

**Support Vector Machine ROC Plots**

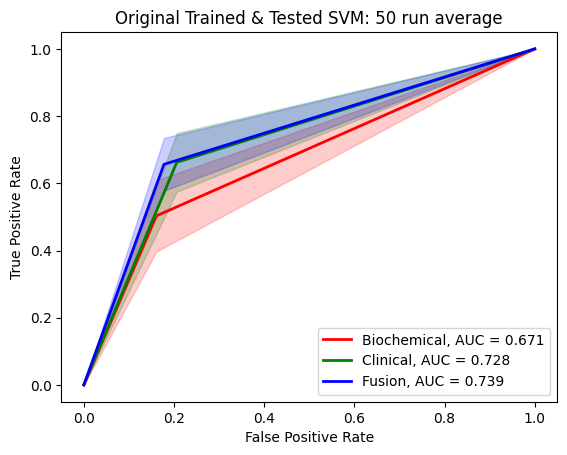

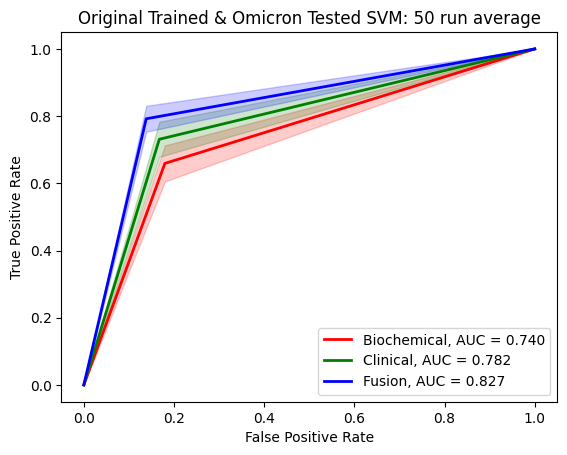

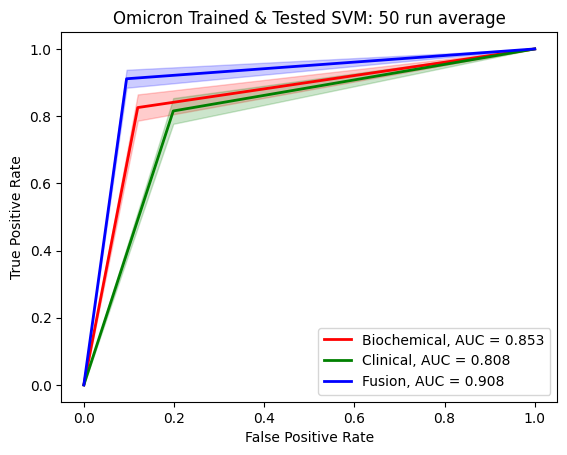

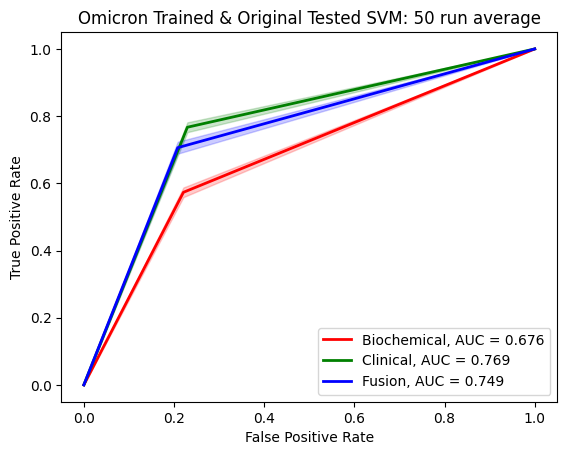
